# Intervention Mapping: A Multi-stage Investigation to Control Gastrointestinal Tract Infections at Primary Level

**DOI:** 10.1101/2025.02.11.25321909

**Authors:** Shahzad Mahmood, Nauman Ali Chaudary, Raja Mohsin Farooq, Hafiz Muhammad Ali Haider, Umer Raza Tahir, Komal Syed

## Abstract

Infectious diseases of the gastrointestinal (GI) tract are among the most common primary healthcare presentations, affecting 2.86 million people worldwide. These infections can include bacterial, viral, parasitic and fungal causes, and may present with nausea, diarrhea, abdominal pain, fever and dehydration. The objective of the study is to decrease disease burden through community transmission mitigation. Three selected key strategies illustrate how the mix-methods approach elaborated the quantitative analysis within the applied qualitative framing. Step 1: Research Phases Pre-Intervention: This research consists of three steps Pre-Intervention, Intervention, and Post-Intervention. In the first phase, disease prevalence is assessed and categorized by age, gender, and geography. The second phase implements a public health intervention tailored to disease risk factors and community needs. The final phase evaluates outcomes through data analysis.Findings highlight that targeted community health measures, such as health education, risk prevention, and intervention mapping, can significantly reduce GI infections, emphasizing the importance of evidence-based public health strategies.

## Introduction

The third world is facing, nowadays, a paradoxical situation of under-expenditure on the health sector and over-production of diseases. An ages-long neglect of preventive healthcare, in both philosophical and practical avenues, is the cornerstone to the dilemma. To address the lack of philosophical backbone, the study is conducted keeping in view the concept of Intervention Mapping (IM) which comprises formation and subsequent implementation of an evidence-based-public health model after the assessment of community needs and acceptability. (1) Community strengthening and stakeholder involvement form the base of the model which encompasses Health Belief Model (HBM), Ecological Theory (ET), and Social Cognitive Theory. Community participation holds a key significance in the research because it guides about the need of particular intervention, from personal to community level, and the mode of intervention and evaluation.(2)

Over the last few decades, the philosophical and practical base of intervention mapping has been strengthened with plenty of articles written on it but some of them address the individual level while the others deal with community level. There is an utmost need for a holistic study that incorporates all aspects of disease prevention, from environmental aspects to personal, social and biological. (3) This study will focus on all those neglected aspects about the spread and containment of gastro-intestinal tract infections. Therefore, the core objective of the study is to design a multiphase research methodology, IM, and then evaluate its effectiveness after its implementation at primary health level. However, the operational objectives include:

- To analyze the prevalence of Gastrointestinal tract infectious diseases in a rural area and segregate them on the basis of age and gender distribution
- To assess multiple factors involved in propagation of GI infectious diseases at primary healthcare level
- To craft and evidence based intervention framework focussing on the control of the spread of infection
- To engage community and stakeholders for behavioral, systematic and environmental change that is vital to reduce the burden of disease
- To analyze the impact of intervention in terms of the change in prevalence of infectious diseases
- To propose a potential and workable mechanism for disease prevention in the form of policy recommendations

The hypothesis thus drawn is, “By Intervention Mapping, healthcare workers can reduce the prevalence of GI tract infectious diseases by 20% after two weeks of effective implementation of a community health program”.

### Ethical Consideration

The research is conducted by giving a key importance to ethics and morality. No participant was forced to join the research and the will of people was central in decision making.The privacy of people was kept intact; their confidentiality was never breached; and no illegality was committed during the whole project. The consent of people was taken before data collection, interviews and in every phase of research a right to consent was primary before taking any step. The study was conducted with a neutral approach by keeping the researchers unbiased and no tampering was made in any sort of data. Succinctly, an ethical approval was taken by ethical review board of the institute of social and cultural studies, University of the Punjab Lahore and the directions by the ethical review board were followed while conducting the research.

## Literature Review

Gastrointestinal tract infections shape a major chunk of infectious diseases at primary level and the stimuli behind their propagation are unhygienic food and poor sanitary conditions. Firstly, Poor water hygiene is the prime source of the disease in rural areas. People do not have access to safe drinking water. Only 20% of people in Pakistan have access to healthy water. (4) The contaminated drinking water is the major source of cholera and diarrheal diseases at basic health unit level. People usually use water from the ponds, canals and ground water for drinking purposes. Generally this water is contaminated by fecal material, bacterias including Escherchia Coli and Vibrio cholerae, drainage and mixing of sewage water, agriculture and industrial waste and animal excreta. The negligence gives rise to a number of water borne infectious diseases including hepatitis, typhoid, dysentery, diarrhea, cholera, and jaundice. (5)

Similarly, food hygiene plays a great role in gastric diseases. In rural areas people are careless in this terrain. Hand washing before meals is obsolete and the food, most of the time, gets contaminated by germs carried through flies, ants, rodents and mosquitoes. Processed food is not packed neatly and frequently it is spoiled. This causes the spread of toxic germs like Salmonella, Shigella, Staph. Aureus and E. Coli, to transfer from one person to another and thus the chain reaction of contagiousness increases transmissibility and infectivity. The frequent cholera outbreaks, as an illustration, in Pakistan coin their root cause from poor food hygiene. (6)

The intensity of issue necessitates the presence of an all-encompassing study which can provide a policy way forward as well as a strong philosophical basis for new research. Intervention mapping is a new terrain of research that finds its exuberant role in public health. It shows new avenues to the researchers. This research project is also conducted by keeping an eye on the ethos of evidence based public health intervention. The six components of intervention mapping are applied systematically in three consequent phases. (7) Program implementation/ Intervention phase (phase 2) is of great importance. It encompasses the fourth theoretical component and forms the bedrock of the study. An organized methodology, consisting of public health programs of health education and awareness, is applied to a population under study. The program forms the breeding ground for preventive medicine and health promotion. (8)

It included periodic demonstrations, health education sessions, public outreach campaigns, health promotion initiatives and disease prevention strategies. The program is implemented by a collaboration of healthcare workers, key gatekeepers, community members and the general population. (9) After a two week exercise of the project, data is collected to interpret a change in prevalence of the infectious diseases after intervention. (10) The results of the study are further elaborated to a gender-particular and age-specific analysis by comparing and contrasting pre and post intervention data.

## Study Design and Methodology

### Research Design

This is a comprehensive intervention design based on the intervention mapping model, addressing various ecological levels, including individual, family, group, and community. Each level is carefully analyzed to identify its specific needs and the corresponding interventions required. The study is structured into three phases, comprising six components.

The first phase, known as the exploratory analysis, involves three sub-levels of intervention mapping. The initial sub-level, termed contextual evaluation and demand analysis, focuses on assessing the disease burden and determining the necessary interventions. The second sub-level is the formulation of program objectives, followed by the third sub-level, which involves the development of program components and design.

The second phase, referred to as the Program Intervention phase, is pivotal and includes a single key component: the Program Adoption and Implementation Plan. Once this phase is successfully completed, the study transitions to the final phase, called the Evaluation and Impact Analysis Phase. This phase comprises two sub-levels: the evaluation plan and the review strategy.

### Study Focus and Sample Population

The study is conducted at a Basic Health Unit in Punjab, Pakistan. It is the primary pillar of primary healthcare in the state. It is a primordial presentation site of patients from nearly 10 villages and coverying 33432 people in a geographical zone of less developed area. The sample conduction site was a general OPD of 24/7 BHU and the intervention site was its field, covering 10 villages in its vicinity. Patients were segregated on the basis of their age and gender and they were asked questions about how they think they were infected and how they think they could have prevented this infection. This was the qualitative component of the study while the quantitative component deals with primary epidemiology of GI infections and the subsequent reduction in their disease load after an intervention.

### Timeline of the Research

The research was carried out in three and half months. The data collection for phase one was initiated on 15th of September 2024 and carried for 45 days till 30 October, 2024. The Intervention phase had a span of 15 days and then the final phase continued from 15 November,

**Table No 1.**
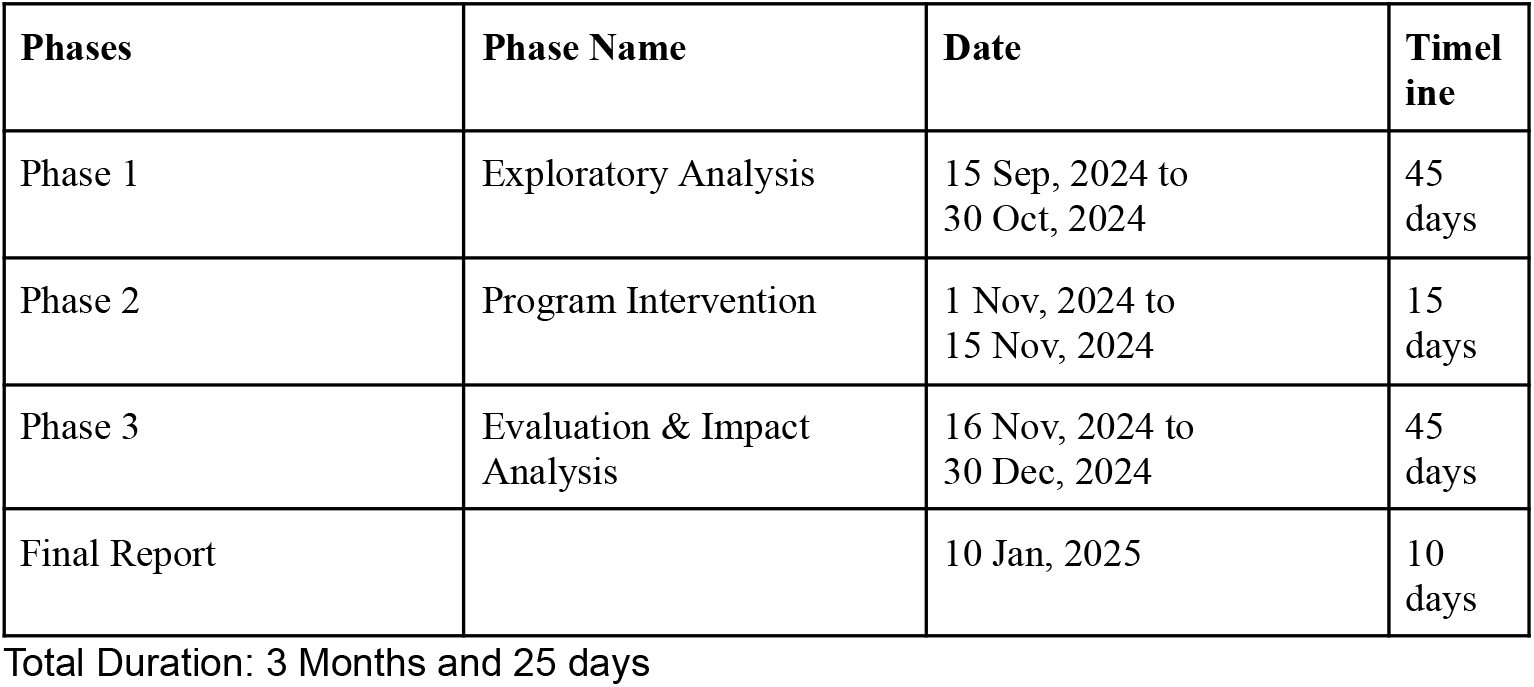
Timeline of Research.

| Phases | Phase Name | Date | Timel<br>ine |
| --- | --- | --- | --- |
| Phase 1 | Exploratory Analysis | 15 Sep, 2024 to<br>30 Oct, 2024 | 45<br>days |
| Phase 2 | Program Intervention | 1 Nov, 2024 to<br>15 Nov, 2024 | 15<br>days |
| Phase 3 | Evaluation & Impact<br>Analysis | 16 Nov, 2024 to<br>30 Dec, 2024 | 45<br>days |
| Final Report |  | 10 Jan, 2025 | 10<br>days |

Total Duration: 3 Months and 25 days

### Exploratory Analysis(Phase 1)

The onset of the first phase marked the start of a new arena in the healthcare sector of the research area. It was commenced on 15th of Sep 2024 with an aim at analyzing the epidemiology of gastrointestinal Tract diseases and to carve a way to reduce their prevalence.

### Contextual Evaluation and Demand Analysis

This deals with analyzing the epidemiology of GI infection in different villages under the surveillance of the basic health unit. The quantitative sub-component of phase 1 focuses on collecting data about the prevalence of GI infectious diseases and their segregation according to age and sex. Data is collected from the general out-patient department of the BHU and the number of patients presented with GI infectious disease is recorded along with the name of their village or vicinity and those were 318 with different age and gender distribution as expressed in Table No. 2.

**Table 2.**
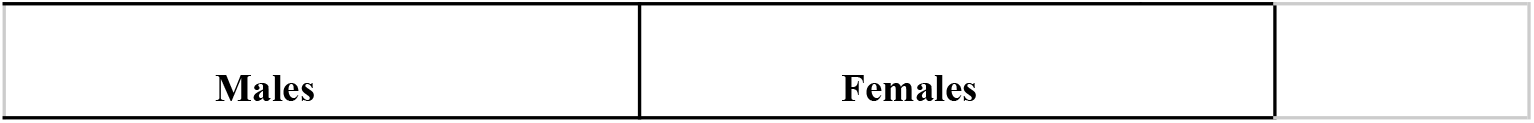
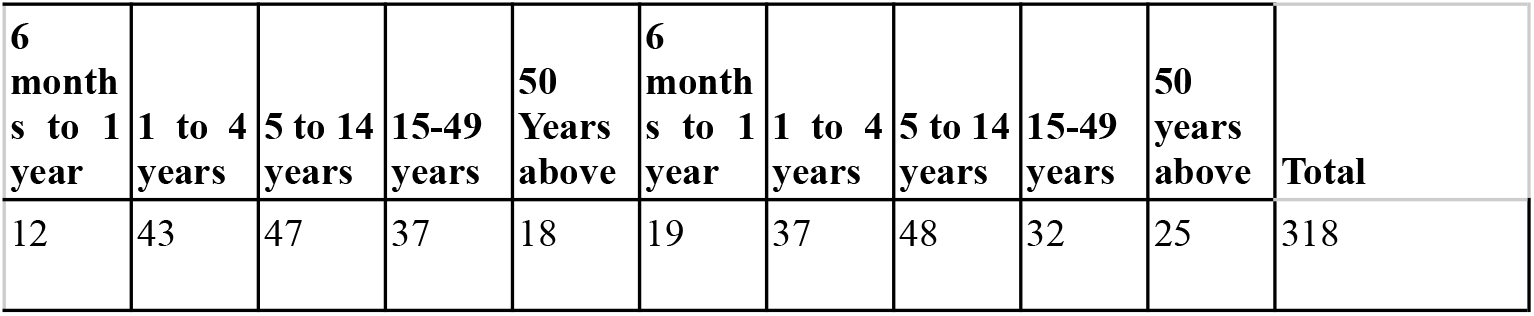
Age & Gender Distribution of Gastrointestinal Infections in Pre-Intervention Phase(Phase 1)

### Production of Program Components & Design

This stage represents the qualitative aspect of intervention mapping. Since the intervention is intended for the community, the primary objective is to gather input from the people to create an effective public health intervention. Data collection in this sub-component of Phase 1 involves structured interviews, community analysis, field surveys, stakeholder perspectives, and ethical considerations.

The primary source of insights was the out-patient department (OPD) of the Basic Health Unit (BHU). Patients visiting the OPD were asked specific questions, such as:

- What do you think caused the disease?
- How do you believe the disease could have been avoided?
- What preventive measures should be implemented in your village to prevent the disease?

In addition to patient responses, community surveys and stakeholder opinions were also highly valued. Key stakeholders included healthcare workers, religious leaders (Imam Masjid), village heads (Lumberdars), community elders, and the Sarpanch. By collaborating with these key community figures, a well-rounded intervention was designed to be implemented during the intervention phase of the research.

### Program Intervention (Phase 2)

Gastric infections are somewhat evenly reported in all age groups so a community approach of intervention was applied to control these infections. The sources of transmission and particular method to contain them were recognized in phase 1. The pre-planned intervention is applied in this phase. Seven GI prevention teams were formed, each consisting of a medical officer, dispenser, lady health workers, lady health supervisor, school health and nutrition officer, and community notables of each village e.g lambardar and imam masjid. Each GI prevention team launched multiple public health advocacy campaigns in its village. The campaigns consisted of seminars, demonstrations, sensitizing lectures, speeches, school & street level campaigns and personal outreach.

The content of health education was very simple. People were taught to use clean and safe water. They were encouraged to boil water before use. Females were advised to cook food properly so that germs in poorly cooked meat may not be a cause of infection. People were empowered to use insect repellents in their houses and they were provided with the knowledge on how the flies can transfer the infectious agents. Furthermore, mothers were advised to pasteurize milk and milk products properly before using. The health advocacy campaign in each village commenced on 1st of september and continued for 15 days till the onset of fruit boding or the final phase.

### Evaluation & Impact Analysis Phase (Phase 3)

The epidemiological statistics of gastrointestinal infection also deserves mentioning while studying infectious diseases at rural level. The number of patients reported with gastrointestinal infection from 15 Nov, 2024 to 30 Dec, 2024 were 262 with age and gender distribution as expressed in table no.3.

**Table No 3.** Age & Gender Distribution of Gastrointestinal Infections in Post-Intervention Phase(Phase 3)

| Males |  |  |  |  | Females |  |  |  |  |  |
| --- | --- | --- | --- | --- | --- | --- | --- | --- | --- | --- |
| 6<br>mont<br>hs to<br>1<br>year | 1 to 4<br>years | 5 to<br>14<br>years | 15-49<br>years | 50<br>Years<br>above | 6<br>mont<br>hs to<br>1<br>year | 1 to 4<br>years | 5 to<br>14<br>years | 15-49<br>years | 50<br>years<br>above | Total |
| 7 | 37 | 24 | 46 | 20 | 9 | 31 | 25 | 44 | 19 | 262 |

### Evaluation of Intervention in Terms of Gastrointestinal Tract Infections

A meticulous observation of the data collected in pre and post intervention phases suggest that the number of GI infections were also reduced due to the community health intervention, as analyzed,

Decrease in the No. of GI Infection Cases = No. of the Cases Reported in Phase 1-No. of the

Cases Reported in Phase 3

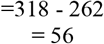

Percentage decrease in the No. of the Cases= (Actual Decrease/Total No. in Phase 1)100

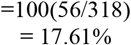

So, a 17.61% reduction in the cases of GI infections has been witnessed. The changing trend is further highlighted in the figure No. 1, where 54.8% of patients were seen in the first phase while 45.2% in the third phase.

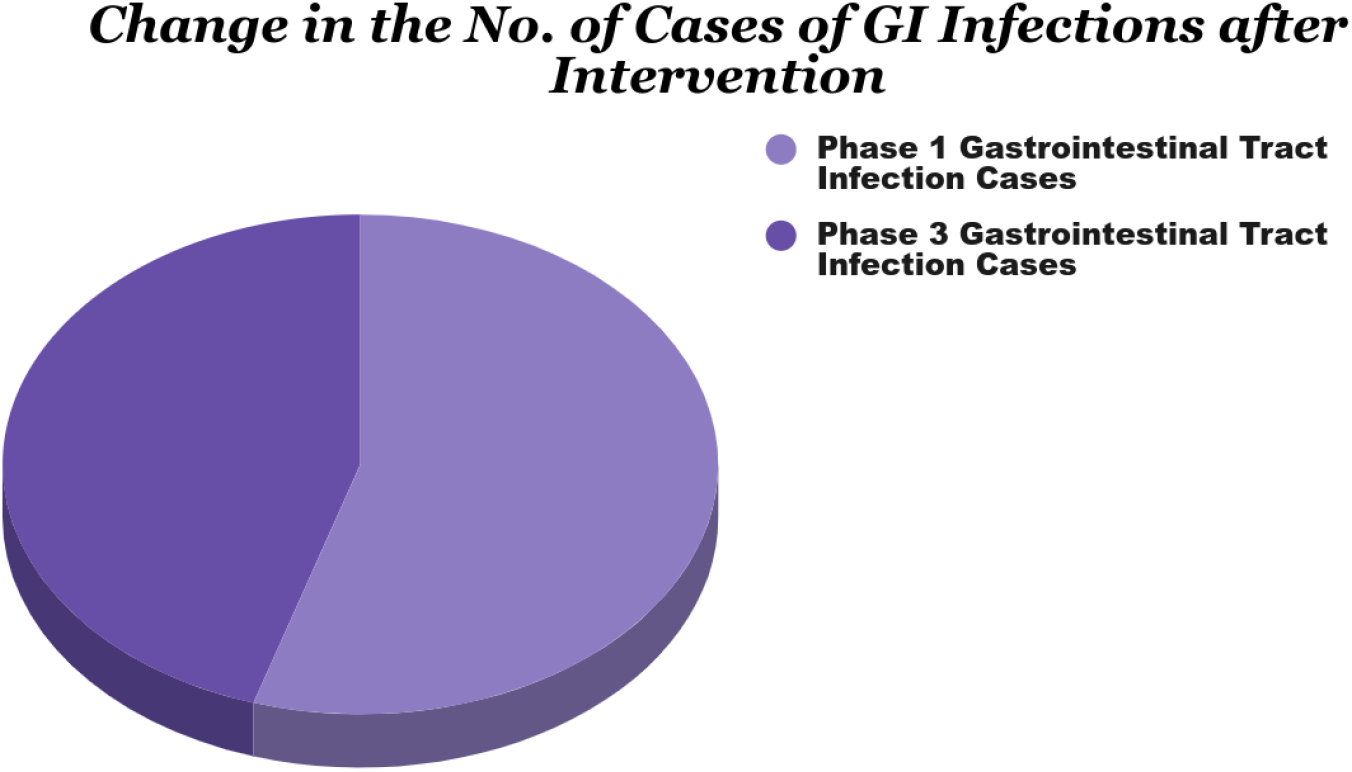

### Age Related Change in Disease Prevalence

The age groups most vulnerable for the disease were targeted specifically and an evidence based public health intervention was made. The data thus obtained in first and last phases showed these trends as described in figure no. 2

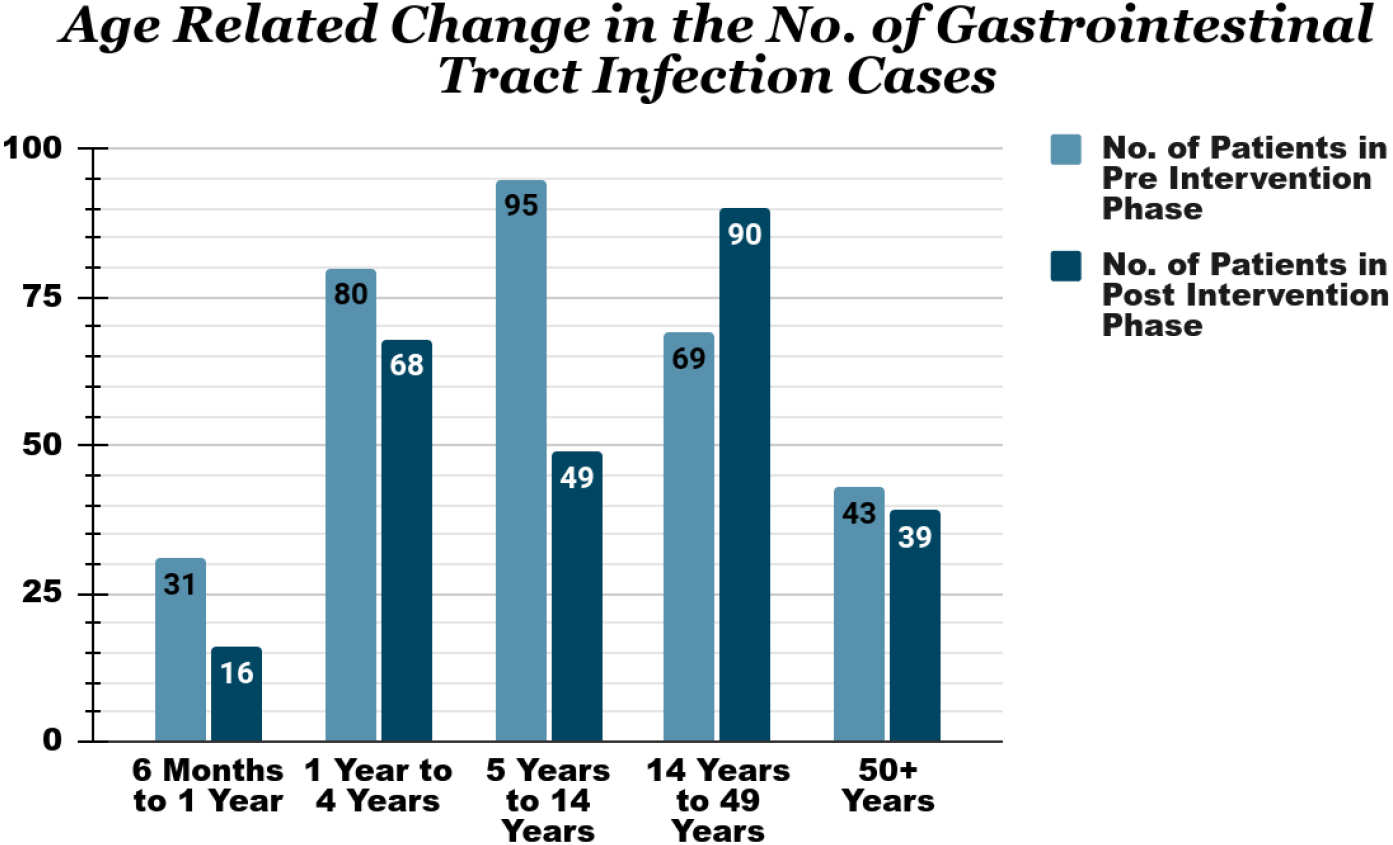

As it is depicted in Figure 2, Age Related Change in the No. Gastrointestinal Tract Infection Cases, the prevalence of GI infectious cases has decreased from 31 cases to 16 cases in the age bracket of 6 months to 1 year, 80 cases to 68 cases in the age bracket from 1 year to 4 years, 95cases to 49 cases in the age bracket of 5 years to 14 years and 43 cases to 39 cases in the age limit of above 50 years. A uniform trend of general decrease in the prevalence of gastrointestinal tract infection after a community intervention was noted except in the people between the age group of 14 years to 49 years, which needs further targeted intervention.

### Gender Based Trends of Gastrointestinal Diseases

The biostatistics of gender based trends are following:

No. of Male Patients Reported with GI Infections in Pre Intervention phase= e1 = 157

No. of Male Patients Reported with GI Infections in Post Intervention phase= f1 = 134

Decrease in No. of Male Patients Reported with GI Infections= z1 = 157 - 134 = 23

Percentage Decrease=Z1 = (e1/f1)100 = (23/157)100 = 14.64%

No. of Female Patients Reported with GI Infections in Pre Intervention phase= e2 = 161

No. of Female Patients Reported with GI Infections in Post Intervention phase= f2 = 128

Decrease in No. of Female Patients Reported with GI Infections= z2 = 161 - 128 = 33

Percentage Decrease= Z2 = (z2/c2e)100 = (33/161)100 = 20.496%

As shown in Figure No. 3, biostatistics depict a general decrease in the patient load among both the sexes after intervention. Nevertheless the reduction is more pronounced in females with 128 female patients reported in the post-intervention phase as compared to 161 female patients in the pre-intervention phase. On the other hand, 134 male patients were reported in the third phase as compared to 157 in the first phase. It clearly shows a reduction in the disease load after a successful public health intervention. While this reduction was more pronounced in females as compared to the males as a 20.496% decrease in patient load in females while in those of males, it was 14.64%.

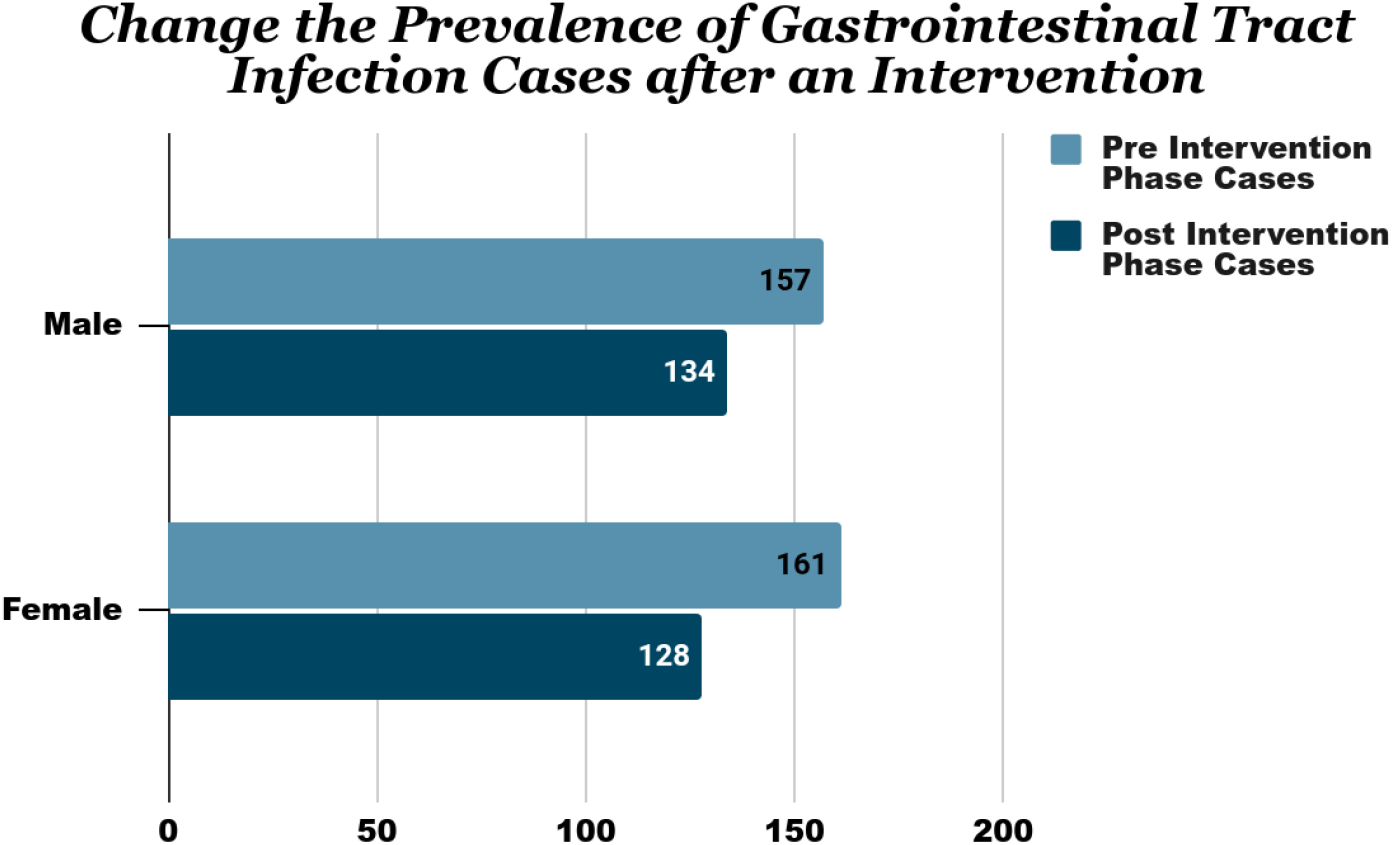

## Conclusion

There is a drop of 17.6% in disease load after the commencement of a community health initiative. All age groups are seen following the general fashion of reduction in disease epidemiology as illustrated: a 15% drop in infectivity of the children (aged between 1 to 4 years); and nine percent decrease in the cases among people above 50 years of age. Adding more, the burden of disease in the age bracket of 5 to 14 years and the infants is reduced to half after a successful public health intervention. It strengthens the assertion that health promotion and disease prevention programs at primary level can deter the spread of gastrointestinal tract infections. (11)

However, a slight conflicting wave from the common fashion is observed among the people of age between 15 to 49 years. A 15.9% rise in the reporting of the number of patients has occurred after the implementation of the population health project. The irreconcilability in this tendency suggests a greater focus on this sub-group is particularly needed to hinder the spread. In Spite of less focus, a change in environment also holds a key impact over this trend. People of rural areas are intricately connected with agriculture and land. Keeping this in view that this was the cotton picking season and people working on fields the whole day usually do not care about their personal hygiene, (12) particularly hand washing before eating, which becomes a source of disease. This explains the absurd trend in the people aged between 15 to 49 years, i.e the age of labor. (13)

The practical implications of the study are clear and evident from its bright outcome. It directs the policy makers to adopt a preventive model at primary healthcare level rather than working on a quasi-curative that is neither preventive nor curative. Each lady health worker, for example, in Pakistan is bound to bring at least two delivery cases to the BHU in one month. It, no doubt, is a step to lessen the sufferings of maternity and improve mother and child health. However, unintentionally, it has become a step to incentivize LHWs to enhance the population of the state, when the whole globe is striving for population reduction. Similarly, this study also highlights the importance of community health besides curative approach. It entails that if disease prevention becomes the core focus, the disease load over the BHUs can be minimized, if not eradicated altogether.

The assertion might seem absurd, but it is not beyond the grasp. If an all-out effort can eradicate smallpox from the world, the dream of rooting out infectious diseases from the globe can also be materialized, but only with joint ventures among healthcare workers, policy makers and community members. The first step that at national level the government can take is to appoint a general physician who is also equipped with a public health degree along with a degree in medicine and surgery needs to be appointed at each basic health unit. The aftermaths of this corollary are bifold. Besides taking care of the community through preventive health care and curative medicine, they will become an instrument for new policy making with research and development at primary level.

## Data Availability

All data produced in the present work are contained in the manuscript

## Notes

### Competing Interest Statement

The authors have declared no competing interest.

### Funding Statement

The study did not receive any funds

### Author Declarations

Ethical approval was taken from the ethical review board of Institute of Social and Cultural Studies, University of the Punjab Lahore, Pakistan.

